# A two-wave epidemiological model of COVID-19 outbreaks using MS-Excel^®^

**DOI:** 10.1101/2020.05.08.20095133

**Authors:** Agenor De Noni, Bernardo Araldi da Silva, Felipe Dal-Pizzol, Luismar Marques Porto

## Abstract

The emergence of the coronavirus SARS-CoV-2 has raised a global issue and a pandemic disease outbreak, COVID-19, was declared by the World Health Organization on March 12^th^, 2020. The new virus is rapidly spreading in humans and cases of severe acute respiratory syndromes are being reported worldwide. Health authority advisors and governments from small towns to large countries need to quickly manage and deal with growing epidemiological data on a daily basis. In this work, current available data from reported cases and deaths over time were analyzed and treated. Lethality has been calculated by finding linearization of death cases against reported ones, using a time-delayed data transposition. A two-wave statistical model, 2WM, based on the superposition of normal distributions was used to fit current data and to estimate the evolution of infections and deaths, using Microsoft^®^ Excel. The model showed good agreement even for apparent single wave behavior in some countries and can easily be extended to any number of waves. A gamma distribution was used as a risk function to estimate death probability from patient admission to reported death. Evolution of fatality cases over time can then be estimated from the model with reasonable accuracy. Data from South Korea, China, Australia, Germany, Italy and Spain were used to validate the model. Data from The United States, United Kingdom and Brazil were used to study the epidemiology as the pandemic progresses. Additionally, the 2WM was applied to world data and to the Brazilian state of Santa Catarina. The model was implemented in MS-Excel, a popular and easy to use analytical tool. A template spreadsheet is provided as supplementary material. Constant lethality can be determined from the initial stage of the pandemic wave. Values ranged from 1.7% to 15.3%, depending on the degree of possible sub notification cases. Even for places with low testing, a linear relationship can be found. The two-wave model can be fine-tuned to properly adjust the data. The second wave pattern was estimated according to the first wave parameter. The accuracy for estimating COVID-19 evolution was compared to the classic SIR model with good agreement. According to the model, based on current trends, health protocols and policies, approximately 10,000,000 cases and 860,000 deaths will be recorded worldwide. Approximately 99% of that number would be reached by the end of July 2020 given constant conditions.

## 1. Introduction

By the end of 2019, the World Health Organization (WHO) [1] noticed that cases of pneumonia from unknown causes were disclosed in Wuhan City, Hubei Province of China. After that, the WHO announced that it was an odd species of coronavirus (2019-nCoV). The novel species was further named by the International Committee on Taxonomy of Viruses as severe acute respiratory syndrome coronavirus 2 (SARS-CoV-2) on February 11^th^, 2020 [2]. It is coming up as the third highly pathogenic disease that rise in the last 20 years [3]. The transmission rate has been heavily described and the number of deaths has been increasing exponentially. By May 3^rd^, 2020, WHO reported 3,356,205 confirmed cases and 238,730 confirmed deaths, spread around 215 countries, areas or territories.

An investigation [4] reported that the virus may be originated from bats and its transmission associated with a seafood market (Huanan Seafood Wholesale Market) in China [5]. COVID-19, the coronavirus disease promoted by SARS-CoV-2, can be compared with other disease outbreaks such as Ebola [6] and Influenza H1N1 [7] which infected and killed a great number of people worldwide, but it increases the pressure on particular demands of health systems around the world, due to ICU and respirators for longer times. Since the initial outbreak of COVID-19 efforts are being made to better understand the syndrome and its agent infectious pattern. The Center of Systems Science and Engineering at Johns Hopkins University claims that the most important way to measure the burden of COVID-19 is mortality. Different fatality ratios have been reported throughout the countries worldwide, which is defined as the number of deaths divided by the number of confirmed cases [8]. Differences in mortality have been attributed to the differences in the number of people tested, demographics, characteristics of the health care system, among others.

The literature discusses the fatality rate, which represents the proportion of cases of who eventually die from a certain disease [8], although it is only possible to calculate it once the epidemic has ended by dividing the number of deaths by the number of cases. When the epidemic is on course, as it is the case of SARS-CoV-2 disease outbreak in the first months of 2020, this formula can be inaccurate. On the other hand, there is urgent need to better understand the spread of the disease and its outcome and risks. Particularly, one wishes to mitigate the pandemic in order to buy precious time, so the health system is better prepared to deal with the incoming patients. Researchers are using different and complementary approaches such as reviewing techniques for disinfection of surfaces [3], studying transmission factors and their dynamics [9], looking for the origin of the virus [4] and developing alternative mathematical models to predict transmissibility and COVID-19 evolution [2]. Due to the easiness and the achievements of good results, mathematical models are being built to estimate the dynamic of the transmission of the virus [1, 4, 6]. However, some models require software only familiar to experts and many parameters are necessary to process and interpret the data. Sometimes those tools are not easily available and/or are not easily accessible for people directly involved with local data management and critical policy decision. Although there is a lack of good data sets, everywhere, mostly due to the small number of tests, the reported data are those which the leaders, health authorities and public agents have in order to support their decisions, trying to balance saving as many lives as they can, at the minimal economic and social costs.

Many of the problems related to the emergent SARS-CoV-2 remain poorly understood and a lot of efforts have been made to overcome those concerns [11]. Therefore, the aim of this work is to contribute to data analysis and treatment providing a simple way to predict possible scenarios for pandemic evolution from a country down to a state or city using the well-known software MS-Excel, based on data with low accuracy but promoting satisfactory and, hopefully, useful results. Instead of using somehow more difficult to handle deterministic models based on a set of differential equations, we have chosen to use more intuitive statistical modeling tools. Statistical approach was also suggested in a recent published paper [9]; in that case the author applied a hierarchical five-parameter logistic model and one wave, solved by a scripting code in R. The aim of this work is to contribute to the SARS-CoV-2 pandemic analysis as it progresses, offering a very simple but useful fitting model based on classical statistics, a combination of gaussian and gamma distributions, implemented in a MS-Excel spreadsheet. In addition, we provide a simple and easy to use MS-Excel (.xlsx) file and a brief tutorial so the interested user can use to easily follow SARS-CoV-2 epidemic progress.

## 2. Methods

### 2.1 General description

The schematic diagram shown in Figure 1 shows the general approach we have used. Total number of new cases are fitted to a two-wave model, the superposition of two normal (gaussian) distribution functions and, in parallel, death cases are linearized against case numbers (confirmed infection cases). A hazard function is used to consider clinical data (hospitalization time) and evolution of death is then calculated. Model adjustment and actual data fittings and validation are continuously updated to treat new data.

**Figure 1.**
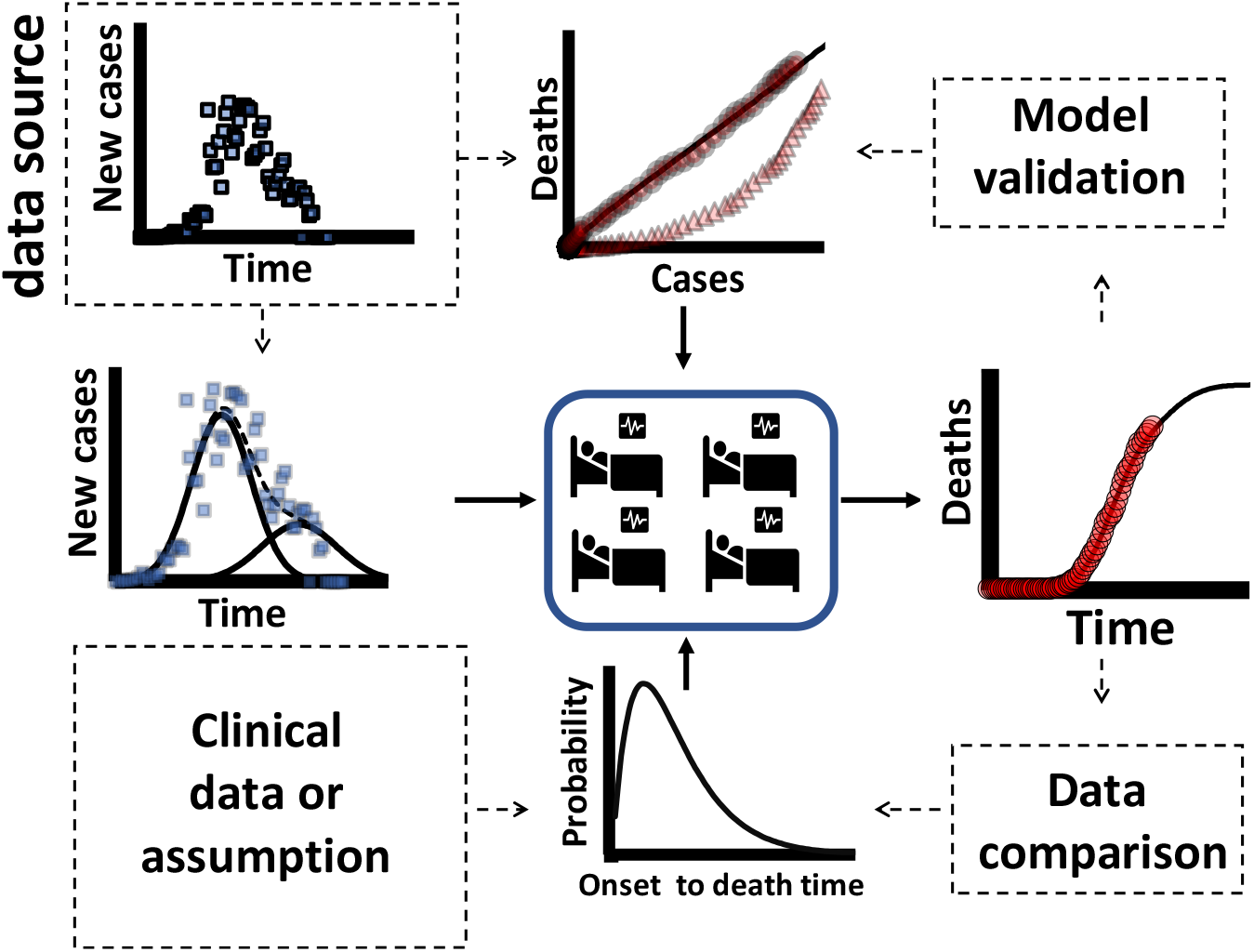
Schematic diagram showing how data source is analyzed and treated. Total number of new cases are fitted to a two-wave model and, in parallel, death cases are linearized against case numbers. A hazard function is used to consider clinical data (hospitalization time) and evolution of death is calculated. Model and actual data adjustment and validation are continuously updated to fit new data.

### 2.2. Modeling

Epidemiological data are being reported as cumulative confirmed cases, active cases, number of deaths and recovered cases. Other relevant parameters are the incidence and testing rates (given in cases per 100,000 people) and the case-fatality and hospitalization rates (usually given as percentage rate). The modeling developed and applied in this study uses the cases and deaths officially reported. In order to treat the epidemiological data as the disease evolves, we have assumed the following hypotheses:

1. Cumulative data behave like a sum of normal, gaussian (normal) distribution, curves;
2. The data were fitted by the addition of two gaussian curves;
3. Each gaussian was named as a single wave;
4. The onset of symptoms corresponds to the beginning of the patient admission date;
5. There is no delay between the date of death and the date of its communication into the public database;
6. A probability of death during hospitalization is implemented as a hazard function, *h*_0_(*t*), and assumed to follow a gamma distribution, i.e., *h*_0_*(t)* = Γ(*X*; *α, β)*, with *α* = 12 *β* = 1;
7. The average onset time is constant throughout the outbreak.

Below we briefly introduce and describe major model variables and parameters.

Model input variables for the gaussian distribution and linearization
*C* Total (cumulative) number of cases (confirmed infections)
*D* Total number of (confirmed) deaths
*X* Time vector (in days)

Model parameter for the gamma distribution
*α* Parameter of gamma function, assumed to be equal to 12
*β* Parameter of gamma function, assumed to be equal to 1

Output variables
*L* Virus lethality, determined by linearization of the observed data from *C* and *D*
*X_i_* Time at day *i (i* = 0, 1, 2,…)
*C(X_i_)* Evolution of total number of cases as a function of time, determined by adjusting the model to the reported data
*D(X_i_)* Prediction of daily deaths as a function of time, calculated from the relationship between *L*, *C(X_i_)* and *Γ*(*X*)

#### Model equations

##### Lethality, L

The lethality of the virus was determined through linearization by translation of the abscissa axis according to Equation 1:

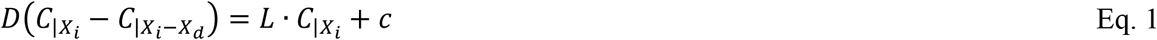

where *X_d_* is the time interval between the date of announcement of the case and the respective death; *c* is the linear coefficient of the equation; *L* and *c* are calculated for the *X_d_* interval in order to maximize R^2^. Usually, *c* value is found to be zero.

##### Cumulative number of cases, *C*(*X_i_*)

The evolution of cases over time, *C(Xi)*, can be described from the superposition of shifted gaussian distribution curves in order to fit the data, according to Equations 2 to 4 of individual waves. In this study, the deconvolution of the cases in two sequential waves was used, but the reader may see that additional cycles could benefit from a greater number of wave outbreaks.

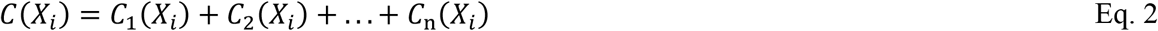

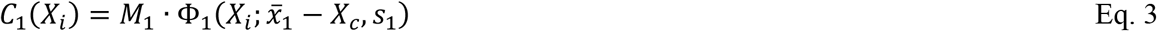

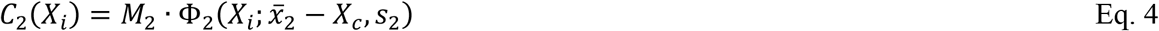

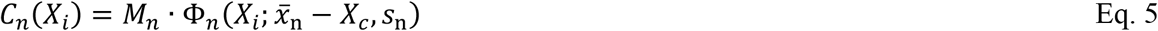

where:

*M*_1,2,…,n_ Total number of cases in the waves 1, 2 and n 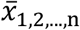 Time at peak of cases in waves 1, 2 and n the average time of the respective cycle *S*_1,2,…,n_ Standard deviation in waves 1, 2 e n *X_c_* The time delay between the date of patient admission and the announcement of a new case.

The parameters *M*_12_, 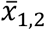, *s*_1,2_ were determined by adjusting the observed data to the model, by least squares, for countries where the outbreak was at advanced stage being possible to identify with significant clarity the behavior of the second wave. *X_c_* is a parameter adjusted from the results of Eq. 6.

##### Cumulative number of deaths, D(Xi)

The prediction of the cumulative number of deaths was calculated from Equation 6.

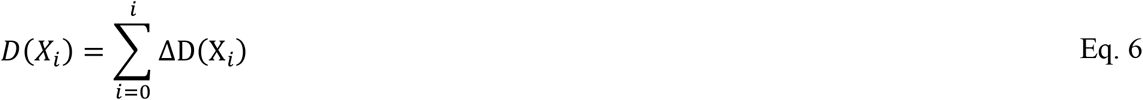

where we have introduced the gamma function as the hazard, probabilistic weighting function, to account for the hospitalization period.

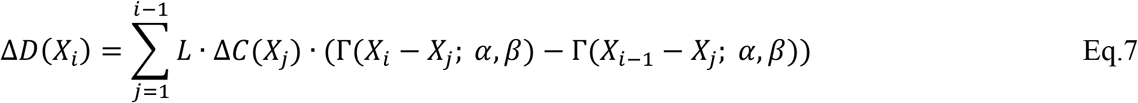

The increase in the number of cases is simply calculated as

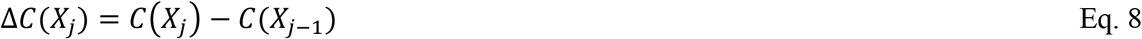

In cases where different lethality is mathematically identified, L_1_ and L_2_, are found for each C_1_ and C_2_ waves, respectively. Equation 9 was used rather than Equation 7 and Equation 10 instead of Equation 8.

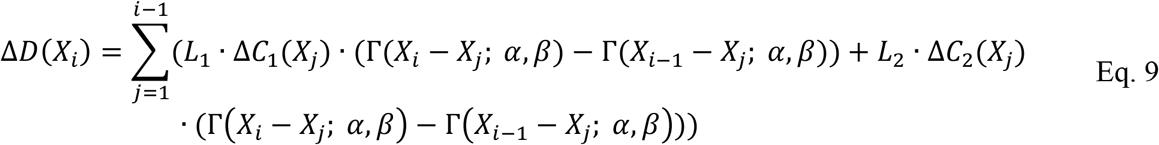

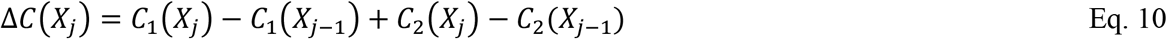

### 3.3. Procedures

Data source regarding cases and deaths through time for selected countries was obtained from Our World in Data [12] with last update in April 24^th^, 2020. South Korea (KOR), China (CHN), Australia (AUS) Germany (DEU), Italy (ITA) and Spain (ESP) were selected as reference countries to test and validate the model. Clinical reported data was applied to reference cases for time distribution from admission to death [13] to fit the gamma function. The major reference was a Chinese study that reported 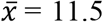 days and *s* = 10.6 days for 40 individuals, ~90% ranged between 4 and 23 days, using α = 2.0 and β = 4.7 [13]. Those parameters were applied to China and South Korea. For other countries where that clinical information could not be obtained the authors have established the equivalent set of α = 12 and β = 1. Therefore, α can directly represent the average time from admission to death, ranging from 4 to 23 days, which represents 99% of death probability.

Microsoft® Excel (2019) (Microsoft Inc., Redmond, WA) was chosen as the spreadsheet used to implement the model. MS-Excel has the advantage of being popular, worldwide used, and support the model with its current tools. The Solver® toolbox, a supplement tool available in MS-Excel distributions, was used to determine the best fit, by minimum square regression of Eq. 3 and Eq. 4, sequentially. The outputs are compared with observed data to check the model adjustment.

Once the validation of the model is accomplished, the second wave parameters *(M*_2_, 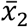*, s*_2_) were expressed as a function of the first wave *(M*_1,_ 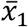, *s*_1_), following the relations given in Equations 11 to 13, below:

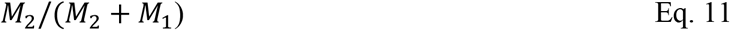

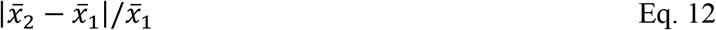

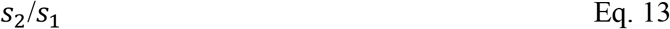

The model was applied to United Kingdom (GBR), United States of America (USA) and Brazil (BRA). In addition, the model was applied to the word (WRLD) and to the state of Santa Catarina (SC), in Brazil.

Results were compared to the prediction calculated by the SIR model with codes available in Matlab® (Mathworks Inc., Natick, MA) [14] applied in April 28^th^ by Singapore University of Technology and Design (SUTD), Data-Driven Innovation Lab, available in the website [15]. The chart data available there was extracted by WebPlotDigitalizer [16]. Comparisons were done using the same data sources (https://ourworldindata.org/coronavirus-source-data), at the same date [12].

## 3. Results and Discussion

### 3.1 Lethality, *L*

Figure 2 shows the number of total current and future deaths against officially registered cases at a certain day for some selected countries as a raw data (RAW = current death) and after linearization (LIN = deaths predicted). In all analyzed cases it was possible to obtain linear adjustment of the data by proper shifting (translation) of the time axis. Linearization procedure was applied to this pair of variables but testing different shifts. It was noticed that each country has its own particularities that this simple procedure can reveal. It was possible to find a lethality value (L) that is constant throughout each outbreak “cycle”. However, for China, South Korea and Australia the procedure revealed two straight line segments. This does not mean that in practice the outbreak had two different lethality. It just shows that conditions may have changed for the disease dynamics or for the data registration protocol, possibly for both. Still, for mathematical modeling purposes, it will be considered that the outbreak may be adjusted as having two pseudo-lethality values, even for cases where those conditions have not changed.

**Figure 2.**
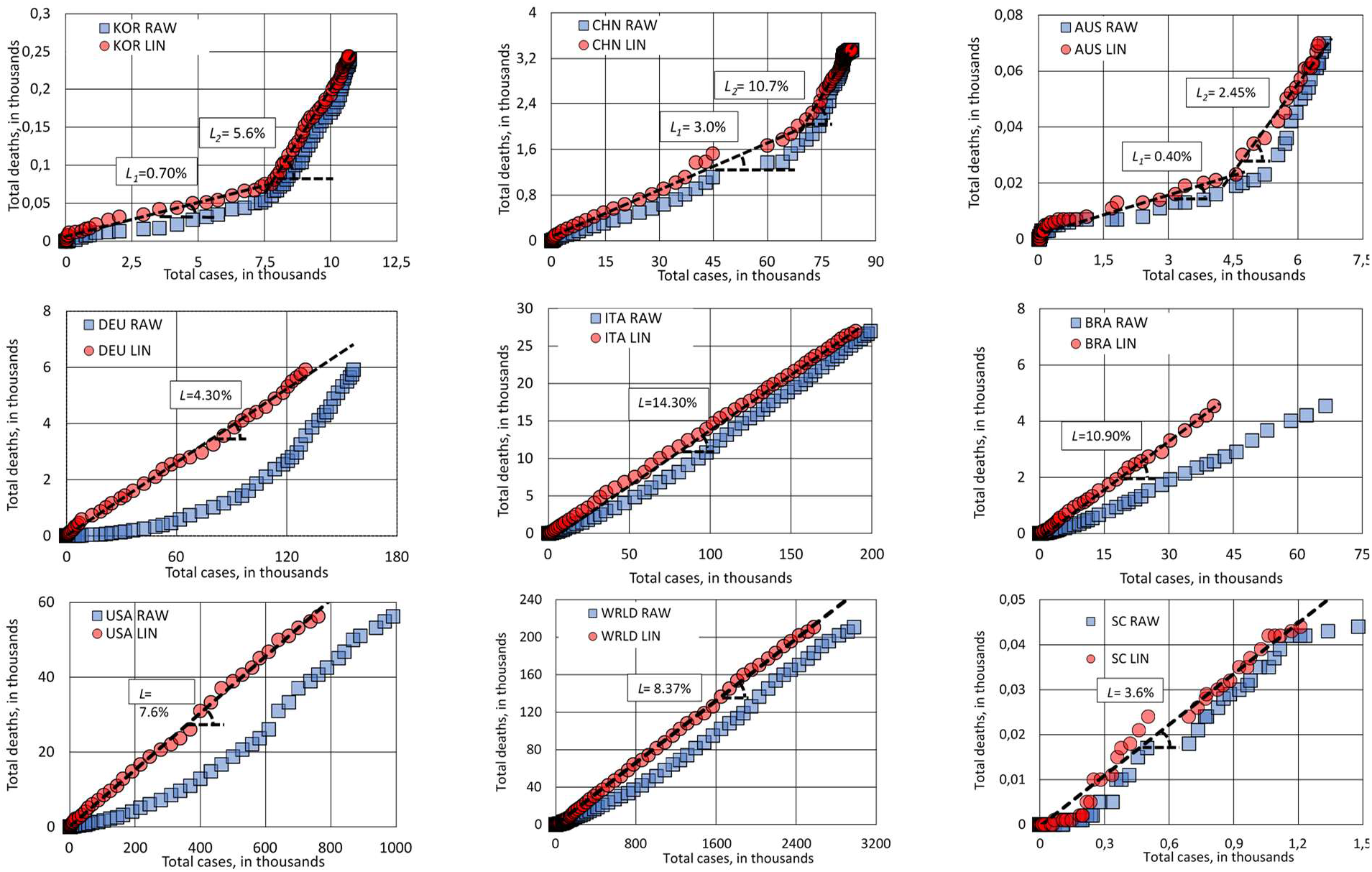
Total number of deaths against total number of cases without linearization (RAW) and after linearization (LIN). This procedure could disclosure a linear relationship between death and cases along the outbreak for a given time shift in one of the variables. The case of KOR, CHN and AUS is detailed in the discussion section.

Table I shows some important information about the pandemic for the selected countries. This table also shows the quantitative results of the linearization procedure, *L* and R^2^. Despite the differences on testing policies between countries, ranged from 1.4 to 27.8 tests per thousand inhabitants, all cases presented a high linearity behavior, high correlation coefficient, R^2^, as well as a low error on *L* values. This result indicates that even with low testing, the information on the number of cases can be linearly associated with the deaths that will be caused by SARS-CoV-2. Thus, even with underreporting of cases, if a country maintains a regular testing policy, it can use those results with as a reliable indicator to measure trends in the evolution of the pandemic. Of course, we know that the absolute values of the lethality may vary. For the countries we have considered, it goes from 1.7% (AUS) to 14.3% (ITA), with a world average of 8.4%. These figures require a more in-depth analysis of the particularities of each country. The problem of underreporting has been widely reported. A country with a high lethality value may indicate that it has a higher rate of unreported cases.

**Table I.**
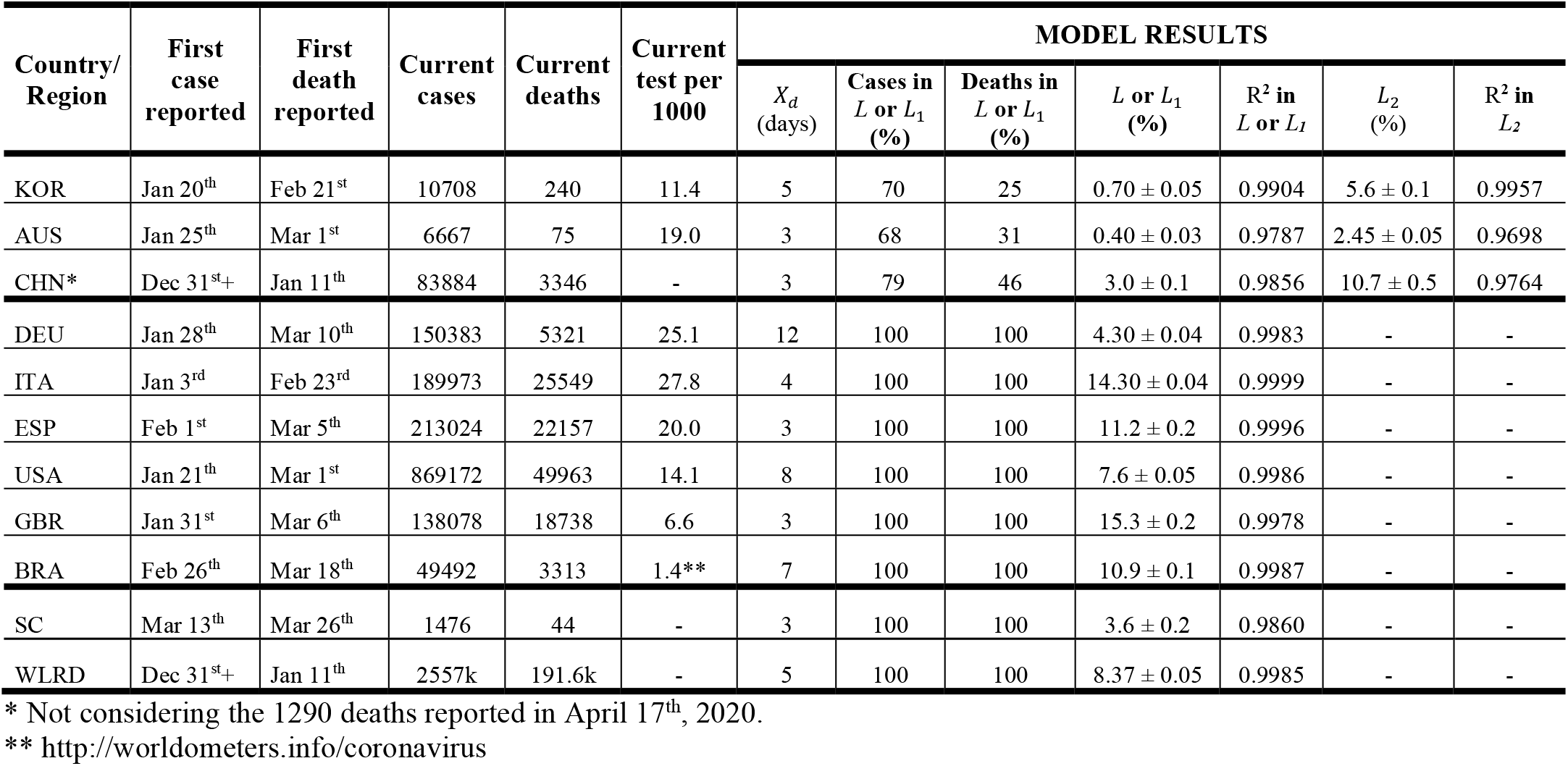
Pandemic data from selected countries or region, lethality and other linearization results as of April 24^th^, 2020 (Source:http://ourworldindata.org)

| Country/<br>Region | First<br>case<br>reported | First<br>death<br>reported | Current<br>cases | Current<br>deaths | Current<br>test per<br>1000 | MODEL RESULTS |  |  |  |  |  |  |
| --- | --- | --- | --- | --- | --- | --- | --- | --- | --- | --- | --- | --- |
| | | | | | | $X_d$<br>(days) | Cases in<br>$L$ or $L_1$<br>(%) | Deaths in<br>$L$ or $L_1$<br>(%) | $L$ or $L_1$<br>(%) | $R^2$ in<br>$L$ or $L_1$ | $L_2$<br>(%) | $R^2$ in<br>$L_2$ |
| KOR | Jan 20<sup>th</sup> | Feb 21<sup>st</sup> | 10708 | 240 | 11.4 | 5 | 70 | 25 | $0.70 \pm 0.05$ | 0.9904 | $5.6 \pm 0.1$ | 0.9957 |
| AUS | Jan 25<sup>th</sup> | Mar 1<sup>st</sup> | 6667 | 75 | 19.0 | 3 | 68 | 31 | $0.40 \pm 0.03$ | 0.9787 | $2.45 \pm 0.05$ | 0.9698 |
| CHN* | Dec 31<sup>st</sup>+ | Jan 11<sup>th</sup> | 83884 | 3346 | - | 3 | 79 | 46 | $3.0 \pm 0.1$ | 0.9856 | $10.7 \pm 0.5$ | 0.9764 |
| DEU | Jan 28<sup>th</sup> | Mar 10<sup>th</sup> | 150383 | 5321 | 25.1 | 12 | 100 | 100 | $4.30 \pm 0.04$ | 0.9983 | - | - |
| ITA | Jan 3<sup>rd</sup> | Feb 23<sup>rd</sup> | 189973 | 25549 | 27.8 | 4 | 100 | 100 | $14.30 \pm 0.04$ | 0.9999 | - | - |
| ESP | Feb 1<sup>st</sup> | Mar 5<sup>th</sup> | 213024 | 22157 | 20.0 | 3 | 100 | 100 | $11.2 \pm 0.2$ | 0.9996 | - | - |
| USA | Jan 21<sup>th</sup> | Mar 1<sup>st</sup> | 869172 | 49963 | 14.1 | 8 | 100 | 100 | $7.6 \pm 0.05$ | 0.9986 | - | - |
| GBR | Jan 31<sup>st</sup> | Mar 6<sup>th</sup> | 138078 | 18738 | 6.6 | 3 | 100 | 100 | $15.3 \pm 0.2$ | 0.9978 | - | - |
| BRA | Feb 26<sup>th</sup> | Mar 18<sup>th</sup> | 49492 | 3313 | 1.4** | 7 | 100 | 100 | $10.9 \pm 0.1$ | 0.9987 | - | - |
| SC | Mar 13<sup>th</sup> | Mar 26<sup>th</sup> | 1476 | 44 | - | 3 | 100 | 100 | $3.6 \pm 0.2$ | 0.9860 | - | - |
| WLRD | Dec 31<sup>st</sup>+ | Jan 11<sup>th</sup> | 2557k | 191.6k | - | 5 | 100 | 100 | $8.37 \pm 0.05$ | 0.9985 | - | - |

\* Not considering the 1290 deaths reported in April 17<sup>th</sup>, 2020.
\*\* http://worldometers.info/coronavirus

Table I also shows the corresponding *X_d_* for each country. Germany has presented the highest value, needing *X_d_ =* 12 days for the linear relationship between death and case numbers. This value match with the average time of 11.5 day from admission to death reported in the literature [13]. We speculate that, in the case of Germany a very systematic procedure between sampling, test result and communication are been conducting. So, the delay time for Germany is *X_c_* ≈ 0. Data from the USA also show typical dynamic delay response with *X_d_* = 8 days. If time from hospitalization to death in the USA is about 12 days, them *X_c_*≈ 4 days. However, it is only possible to estimate an accurate average value of *X_c_* for a particular country or region if the mean admission to death time is known. Considering the information from the literature [13] as aleatory sampling, a confidence interval could be estimated for the average time from admission to death as 9 to 15 days.

### 4.2. Case evolution, *C*(*X_i_*), and Δ*C*(*X_j_*)

Figure 3 shows the graphs of new cases over time for the analyzed countries. The two-wave model fitted the cases satisfactorily. KOR is the only country where the observed data clearly shows a true second wave. In other countries, deconvolution by the two-wave model suggests the possible influence of a second wave, superposed on the previous one while was still progressing. KOR, CHI and AUS outbreaks apparently had a break and the two-wave model was sufficient to describe the cases so far. DEU, ESP and ITA outbreaks are still ongoing; the two-wave model can then describe data evolution for a near future. It is not possible to state that just two waves will be sufficient to describe all outbreaks around the world for the whole pandemic, but as mentioned before, one can easily add new wave cycles as necessary. Please notice that, in the case of CHN, the new cases announced on February 14th and 15th were not considered in the model. The strong discontinuity observed and reported in the literature [9] is a serious burden for any modeling attempted.

**Figure 3.**
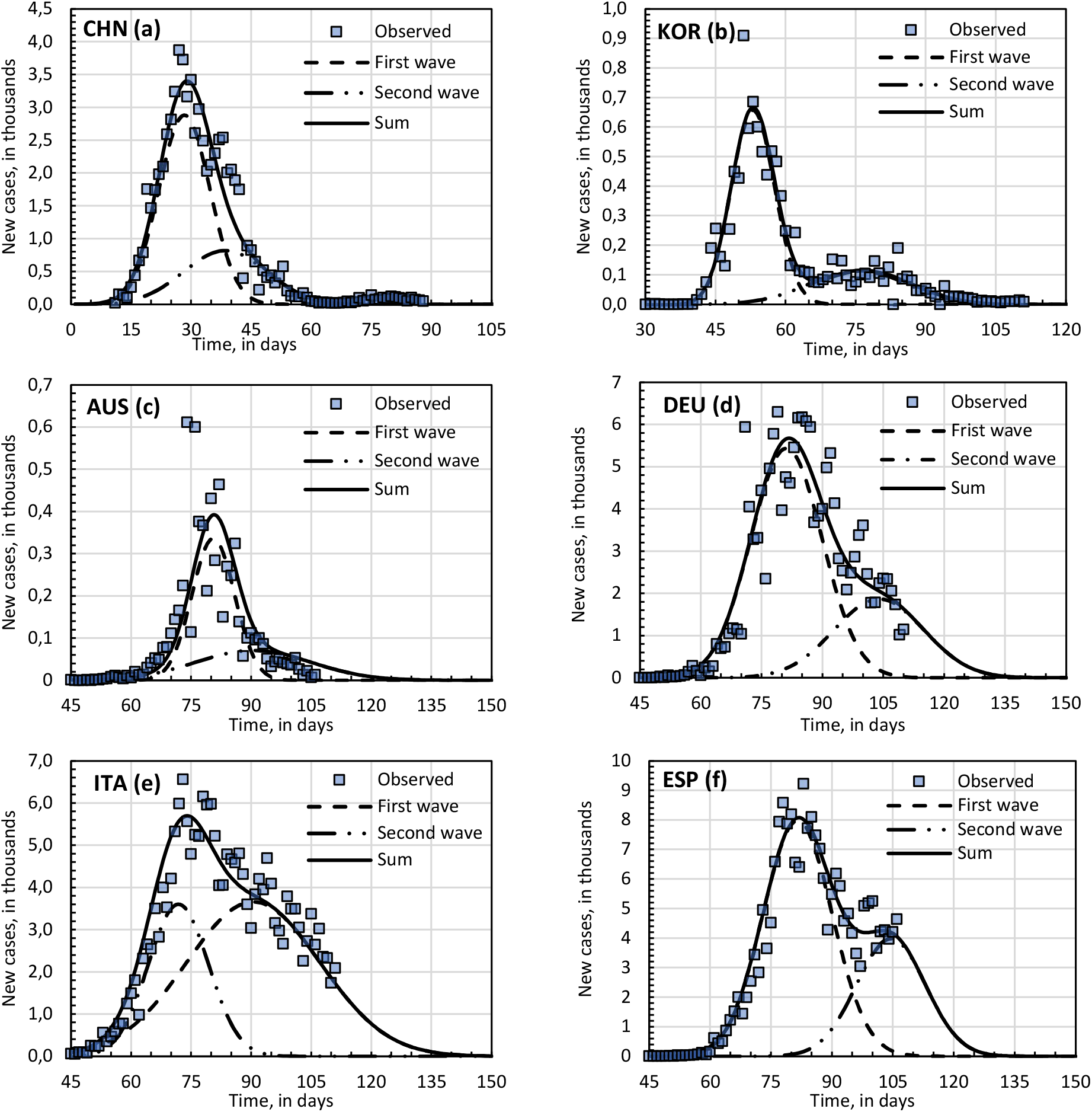
Observed and modeled data for new cases against time. The contribution of each wave to the total effect is detailed. The time zero for all charts is Jan 9^th^, 2020. The two-wave model could satisfactory describe the observed data, considering intrinsic data dispersion.

Table II shows the adjusted parameters found for the reference/chosen countries as well as the parameters for the two-wave model applied to other regions. Another possibility is to use the most expected value for each parameter directly. The standard deviation of the first wave is the key factor for a more accurate prediction, specially at the beginning of the outbreak. It is also important to compare 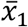 with other countries. Within the error margins it is possible to draw different scenarios and compare different countries/regions. The fitted values for 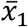 and *s*_1_ for USA, GRB, BRA and SC state were within the limits estimated for the countries taken as reference.

**Table II.** Two-wave model parameters for the reference countries and for the selected countries/regions where the model was applied

| Parameter/<br>Country/region | $M_1$ | $\bar{x}_1$<br>(day) | $s_1$<br>(day) | $M_2$ | $\bar{x}_2$<br>(day) | $s_2$<br>(day) | $M_2/(M_2 + M_1)$ | $\vert \bar{x}_2 - \bar{x}_1\vert /\bar{x}_1$ | $s_2/s_1$ |
| --- | --- | --- | --- | --- | --- | --- | --- | --- | --- |
| Reference countries |  |  |  |  |  |  |  |  |  |
| KOR | 7645 | 42 | 4.6 | 3012 | 65 | 10.4 | 0.28 | 0.57 | 2.25 |
| AUS | 4482 | 61 | 5.3 | 2557 | 71 | 14.37 | 0.36 | 0.16 | 2.71 |
| CHN | 46234 | 37 | 6.4 | 21382 | 47 | 10.4 | 0.32 | 0.28 | 1.62 |
| DEU | 120000 | 63 | 8.8 | 52000 | 85 | 11.1 | 0.30 | 0.36 | 1.26 |
| ITA | 153214 | 70 | 16.7 | 67787 | 51 | 7.5 | 0.31 | 0.27 | 0.45 |
| ESP | 171089 | 59 | 8.5 | 85323 | 81 | 8.5 | 0.33 | 0.37 | 0.86 |
| Selected countries/regions |  |  |  |  |  |  |  |  |  |
| USA | 10660k | 82 | 12.3 | 309 | 109 | 9.9 | 0.32 | 0.35 | 0.8 |
| GBR | 179236 | 74 | 12.6 | 73209 | 100 | 10.1 | 0.29 | 0.35 | 0.8 |
| BRA | 99378 | 59 | 15.0 | 46766 | 97 | 24.2 | 0.32 | 0.35 | 1.6 |
| SC | 1547 | 32 | 13.0 | 495 | 57 | 10.4 | 0.32 | 0.50 | 1.6 |
| WRLD* | 7374k | 107 | 17.2 | 2073k | 157 | 17.9 | 0.32 | 0.5 | 1.04 |

Table III gives first and second wave parameters as calculated from reference countries, i.e., countries chosen to validate the model. The average and range between lower and upper limits may be used to build possible scenarios for a target country or region.

**Table III.** First and second wave parameters as extracted from regression fitting of Eq. 2.

| Wave Parameter | $\bar{x}_1$<br>(days) | $s_1$ | $M_2/(M_2 + M_1)$ | $\vert \bar{x}_2 - \bar{x}_1\vert /\bar{x}_1$ | $s_2/s_1$ |
| --- | --- | --- | --- | --- | --- |
| Average | 55 | 8.0 | 0.32 | 0.35 | 1.6 |
| Lower limit | 37 | 4.6 | 0.28 | 0.16 | 0.5 |
| Upper limit | 70 | 16.7 | 0.36 | 0.57 | 2.7 |

### 3.3. Deaths Evolution, *D(X_i_)*

Figure 4 shows the case and death evolution over the time and deaths as a function of cases for the simulated linearized function. KOR and DEU were selected as representative cases. The combination of two-wave modeling for the cases evolution, lethality and hazard function could estimate the death with a good level of proximity.

**Figure 4.**
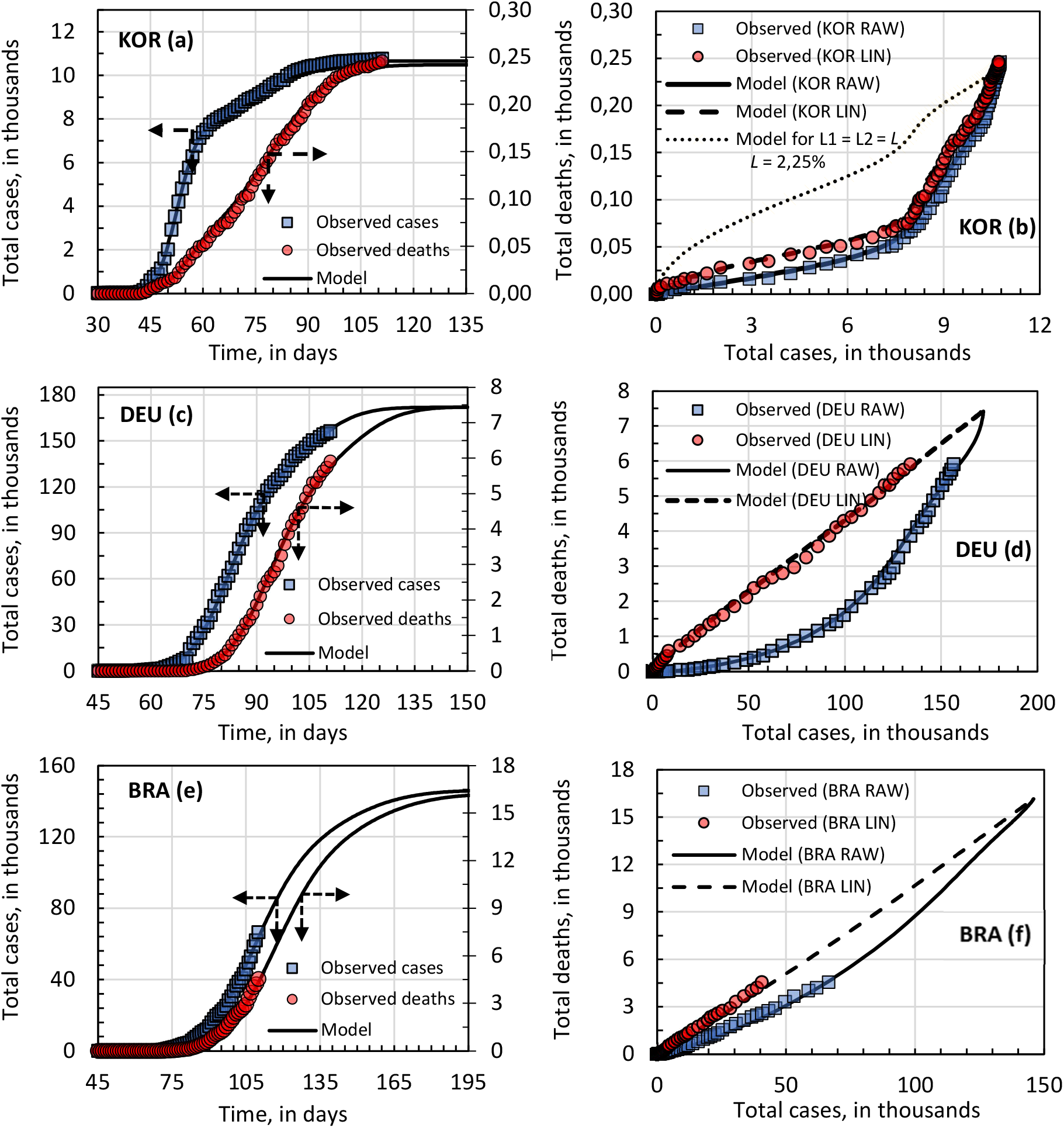
Cases and deaths evolution over the time and deaths against cases for South Korea (KOR) and Germany (DEU), the chosen representative countries. Deaths were calculated combining the two-wave model for cases, lethality, and hazard function. Number of total cases are given in thousands.

South Korea, as well as China and Australia, presented two values for lethality, according to the linearization procedure. Mathematically, this information was not incorporated into the model. However, before concluding that there are in fact two distinct lethality, it is necessary to analyze at least two important aspects: (1) if the duration of the outbreak is faster than the hospital stay until death and (2) if there was any significant change in testing policy or protocol and confirmed cases. The analysis of the data treated here allows to evaluate the first aspect. These countries were the ones that presented the shortest time for the first wave cycle. In the case of KOR the value was *s*_1_ = 4.6 days, see Table II. This means that approximately 85% of people infected in the first wave were infected within 15 days. The average length of stay according to the literature varies from 2 to 56 days, with an average of 11.5 days and standard deviation of 10 days. Thus, it can be said that the period of the outbreak is faster than the length of hospital stays until death. Thus, the lethality of the virus in these countries is more likely to be approximately constant over the outbreak interval. The model would be able to simulate this scenario, but in this case, it would be necessary to obtain accurate information on the distribution of the length of hospital stay until death in the countries considered.

Table IV presents a critical parameter for adjusting the model, *X_c_*, that corresponds to the average delay time between hospitalization and confirmation of the case. The sum *X_c_* + *X_d_* represents approximately the average length of stay until death. If the length of hospital stay is different from that considered in the calculations, the difference may be subtracted from the value of *X_c_*. This adjustment is a position adjustment. If there are no significant variations in the testing protocols during the pandemic, this is a value that does not tend to change. The table also presents a comparison between the predictions obtained with the two-wave model, 2WM, and the SIR model. Differences in the forecast of the total number of cases fluctuated by approximately 20%. The dates predicted to end the outbreak in question also resulted in close values. The biggest discrepancy is related to the forecast for the end of the pandemic in the world. It is observed that the statistical model of two waves allows to obtain forecasts with results similar to those of the SIR, but with the advantages of simplicity and ease of implementation and analysis.

**Table IV.** Estimates for deaths, cases and end dates. Comparison between 2WM and SIR cases. SIR results from: Singapore University of Technology and Design (SUTD), Data-Driven Innovation Lab[15]

| Country/<br>region | $X_c$<br>(day)<br>2WM | Estimated<br>Deaths<br>(thousands)<br>2WM | Estimated Cases<br>(thousands) | | 97% of est'd cases | | 99% of est'd cases | |
| --- | --- | --- | --- | --- | --- | --- | --- | --- |
|  |  |  | 2WM | SIR | 2WM | SIR | 2WM | SIR |
| DEU | 1 | 7.5 | 171 | 230 | May 6<sup>th</sup> | May 4<sup>th</sup> | May 12<sup>nd</sup> | May 16<sup>th</sup> |
| ITA | 8 | 32 | 221 | 280 | May 7<sup>th</sup> | May 14<sup>th</sup> | May 09<sup>th</sup> | May 17<sup>th</sup> |
| ESP | 10 | 28 | 255 | 306 | May 05<sup>th</sup> | May 02<sup>th</sup> | May 09<sup>th</sup> | May 13<sup>th</sup> |
| USA | 5 | 110 | 1440 | 1200 | May 19<sup>th</sup> | May 16<sup>th</sup> | May 25<sup>th</sup> | May 29<sup>th</sup> |
| GBR | 9 | 39 | 252 | 247 | May 22<sup>nd</sup> | May 18<sup>th</sup> | May 28<sup>th</sup> | May 31<sup>st</sup> |
| BRA | 5 | 16 | 146 | 154 | Jun 26<sup>th</sup> | Jun 06<sup>th</sup> | Jul 8<sup>th</sup> | Jun 18<sup>th</sup> |
| SC | 10 | 85 | 2.2 | - | May 22<sup>nd</sup> | - | May 28<sup>th</sup> | - |
| WRLD* | 8 | 860 | 10300 | 11300 | Jul 15<sup>th</sup> | Jun 12<sup>nd</sup> | Jul 26<sup>th</sup> | Jun 25<sup>th</sup> |

## 4. Conclusions

A two-wave statistical model, 2WM, based on the superposition of normal distributions was developed and used to fit current data and to estimate the evolution of infections and deaths in chosen reference countries and other ongoing places. The model showed good agreement even for apparent single wave behavior in some countries and can easily be extended to any number of waves. A gamma distribution was used to estimate death probability from patient admission at a health service to his/her reported death. Evolution of fatality cases over time is estimated from the model with reasonable accuracy. The model was successfully implemented in MS-Excel®, a popular and easy to use analytical tool. Constant lethality was determined from the initial stage of the pandemic wave. Values ranged from 1.7% to 15.3%, depending on the degree of possible sub notification cases. Even for places with low testing, a linear relationship could be found, by properly translating time series data. The two-wave model can be fine-tuned to properly adjust a variety of situations. A second wave pattern was estimated according to the first wave parameters. Confirmation of the future scenario, as predicted by the model, is vulnerable to changes in behavior on the part of the population and policies to deal with the epidemic. As a result, the characteristics of the second wave can extrapolate, for more or less, the observed and parameterized behavior. The accuracy for estimating COVID-19 evolution was compared to the classic SIR model, based on ordinary differential equations, with good agreement. According to our two-wave model and based on current trends, health protocols and policies, approximately 10,000,000 cases and 860,000 deaths will be recorded worldwide by the end of the pandemic. Approximately 99% of that number would be reached by the end of July 2020, given current conditions. In this way, the model offers complementary information to the classic models, hopefully contributing to the monitoring and management of the pandemic.

## Data Availability

All data are were obtained from public access database

http://ourworldindata.org

## Acknowledgements

The authors acknowledge the Brazilian agencies Coordination for the improvement of Higher Education Personnel (CAPES) and the National Council for Scientific and Technological Development (CNPq) for their continuous support.

